# Non-Inferiority Margins in Randomized Controlled Trials in Abdominal Surgery – a Systematic Review

**DOI:** 10.64898/2026.08.29.26361719

**Authors:** Carl-Stephan Leonhardt, Dominique Birrer, Max F. Stauffer, Johannes Toti, Iain J Gallagher, Richard J. E. Skipworth, Barry Laird, Christoph Kuemmerli

## Abstract

**Importance:** Non-inferiority trials are becoming increasingly popular in abdominal surgery. The non-inferiority margin is critical in the interpretation and conclusion of these trials.

**Objective:** This systematic review aims to assess the methodological and reporting quality of non-inferiority randomized controlled trials in abdominal surgery.

**Evidence Review:** Non-inferiority trials were systematically identified by searching Ovid Medline, Embase and the CENTRAL databases from 2006 until December 2025. Randomized controlled trials in adult patients with any type of abdominal surgical intervention in at least one trial arm and a sample size ≥100 were eligible for inclusion. The primary outcome was the definition of the non-inferiority margin. Secondary outcomes were the reporting of the non-inferiority margin, the robustness of its estimation, the uncertainty of the point estimate and the adequacy of conclusions.

**Findings:** A total of 11’045 trials were identified, of which 101 were eligible, enrolling 44’370 patients. Most trials provided a rationale for the non-inferiority design, while six (5.9%) trials did not. Previous literature was commonly used (n=56; 55.4%), but the non-inferiority margin was most often based on a clinical fixed margin or on historical comparison of the treatment and the active comparator. Based on the margin, investigators tolerated substantially worse outcomes of the treatment compared to the comparator.

Conclusions were appropriate based on the confidence interval and the predefined non-inferiority margin in 88 (87.1%) of trials. The clinical judgement of the conclusion was overall adequate. Confidence interval estimations were reported in 16 (15.8%) of trials. Simulation studies were limited by the reporting quality.

**Conclusions and Relevance:** Clinical fixed margins are commonly used in abdominal surgery non-inferiority randomized controlled trials, however, substantial shortcomings in reporting limit the interpretability and reproduction of study findings. Based on the findings of this study, guidance on surgical-specific non-inferiority margin definitions is needed.

**KEY POINTS:** *Question:* How are non-inferiority margins estimated in surgical randomized controlled trials?

*Findings:* In this systematic review, non-inferiority margins were nearly always numerically defined, but reporting of how they were estimated was poor. Most frequently, clinical fixed margins were utilized and previous evidence was either ignored or not included at the design stage. The selected margins often tolerated substantially worse outcomes in the treatment group compared to the comparator.

*Meaning:* Improved reporting and guidance on the conduct of surgical non-inferiority trials are required.

## INTRODUCTION

A non-inferiority (NI) randomized controlled trial (RCT) investigates whether the efficacy of an intervention is not substantially worse than that of an established treatment, also known as the active comparator. NI trials are increasingly used because it is unethical to deny treatments that were proven to be effective in former superiority trials. Hence, the new treatment must offer other benefits, e.g. it is less costly, has a higher availability, the application is easier and/or the compliance is higher, it is less invasive, has fewer adverse events or similar.^1^

The specification of the NI margin is critical in the design of an NI trial. The NI margin is used to assess whether an intervention will preserve a clinically relevant fraction of the effect of the active comparator. The NI trial is powered to demonstrate that the lower bound of the confidence interval (CI) excludes the NI margin. Hence, the NI margin is crucial for the sample size calculation, the interpretation of the trial and therefore, the conclusion derived from the trial findings. While less considered, the method for CI calculation is thus closely related in determining non-inferiority compared to the active comparator.

Various methods exist to define the NI margin, primarily based on clinical and statistical reasoning. For pharmacological trials, the Food and Drug Administration (FDA) and European Medicines Agency (EMA) provide guidance on specification of the NI margin. In contrast, for non-pharmacological trials like surgical interventions, there is little guidance on the choice of the NI margin.^2,3^

Despite efforts to improve the reporting of NI trials - including the development of a CONSORT extension specifically for NI trials – systematic reviews indicate that frequently neither the NI margin itself nor the method used to determine it are reported in pharmacological trials.^4–12^ Similarly, among 347 surgical trials across various specialties and surgical as well as non-surgical interventions, only 28.5% reported on the method of NI estimation.^13^

In addition to poor reporting, non-adherence, missing data, and non-constancy of the treatment effect may blur the interpretability of NI trial findings compared to the more intuitive superiority trials. Notably, an NI design has been shown to lead to outcomes supporting the investigators hypothesis in most cases.^14,15^

The primary objective of our work was to systematically assess the methods used to define the NI margin in abdominal surgery RCTs. The secondary objectives include the reporting quality of these trials based on the CONSORT statement, the robustness of the NI margin estimation, the uncertainty of the point estimate and the clinical adequacy of conclusions.

## METHODS

This work followed the reporting guidance of the Preferred Reporting Items for Systematic Reviews and Meta-Analyses (PRISMA) guidelines whenever applicable.^16^ This work was conducted according to a previously published and preregistered study protocol on the Open Science Framework (OSF) (https://osf.io/3gx4b/).^17^ Post hoc deviations from the protocol are considered exploratory and are clearly indicated.

### Eligibility criteria

To be included, trials met all of the following criteria: (i) RCT with an NI design as specified in the title or abstract, (ii) examines an abdominal surgical intervention, defined as a surgical procedure (e.g. appendectomy or cholecystectomy), or a novel surgical technique (e.g. open or laparoscopic access), (iii) has at least two arms of which at least one must be a surgical treatment, (iv) reports the results of the primary endpoint (PE) analysis for which the NI margin was estimated, (v) was published as full-text from 1st August 2006, (vi) has a trial sample size for the PE analysis of ≥100 randomized participants and (vii) includes an adult study population (participants ≥18 years old). Exclusion criteria were: (i) comparison of medicines, (ii) only endoscopic procedures, (iii) psychological or behavioral interventions, and (iv) PE not reported.

### Information Sources and Study Selection

Ovid Medline, Embase and The Cochrane Central Register of Controlled Trials (CENTRAL) were systematically searched on 20th December 2025.^18^ The choice of the three databases was considered sufficient because our study focused was on randomized trials. A librarian provided support for the choice of the databases and advised on the search strategy. Clinical trial registries were not searched separately as only published trial findings were eligible. Corresponding authors were not contacted for further information.

All original articles reporting the PE of RCTs were searched. For the included studies, additional information sources were identified and searched, including the published trial protocol, the statistical analysis plan, other published result reports (e.g. short-term results) and trial registries. All publicly available sources were considered before a final data point was extracted. Only after considering all sources and no pertinent information was found; a data point was set as “not reported”.

Covidence (Veritas Health Innovation, Melbourne, Australia) was used to import findings from all database searches.^19^ Duplicates were identified automatically, but a sample of 25% of articles was checked manually. The screening was hierarchical in the following order: title, abstracts, full-text article. Based on prescreening, duplicates, non-surgical studies and trials with <100 participants were excluded by one reviewer (CK). Two reviewers screened all remaining articles independently. Disagreements were resolved by a third reviewer. Full-text articles were again screened by at least two reviewers independently after exporting them to Endnote^TM^, version 20.^20^ Finally, RCTs including abdominal surgical procedures were selected.

### Search strategy

Separate search strategies were developed for all three databases (eTable 1). The search string had three components:

i. The study design RCT. The string developed by the Scottish Intercollegiate Guidelines Network (SIGN), which is pre-tested, was used after a slight modification by removing the specification of the phase of the trial.^21^
ii. The NI design, which comprised the following string: non-inferior* OR noninferior* OR NI[tiab] OR equivalence OR equivalent. Equivalence trials were also searched for because this term is sometimes used instead for NI trials.
iii. The surgical specialty. Terms for surgical procedures were introduced to capture all surgical trials. To narrow the search, common terms for surgical specialties and surgical procedures were identified based on MeSH terms.

### Data extraction

The following baseline features were extracted: name of the first author or the collaborative, year of publication, journal name, funding sources and the sponsor of the trial. Further, where the design was specified, the intervention and the active comparator or reference treatment were captured and the phase as explicitly stated by the authors and the blinding procedure, if applicable, were identified. The randomization procedure, sample size calculations, the number of interim analyses, and the PE were also extracted.

Information based on the CONSORT extension was collected. The reporting of the PE was considered of good quality if the results per group, the intergroup difference and the CI of that difference were reported. If there was insufficient information for the interpretation of the findings, i.e. no CI, the author’s conclusion was defined as not appropriate. The conclusion and the clinical judgment of the risks and benefits of the intervention was assessed, regardless of the reporting quality. If the calculated CI crossed the NI margin, non-inferiority was considered rejected or inconclusive and vice versa. If the authors stated otherwise, this was considered an inappropriate conclusion. If the intervention arm performed better than the active comparator, a comparison of the effect with historical evidence would be appropriate to assess whether the constancy assumption was violated. Whether this was assessed was recorded.

### Outcomes

Lack of reporting of a variable and the percentage of unreported data points for all variables based on the CONSORT extension was recorded.

For the definition of the NI margin, historical evidence and statistical considerations were considered as potential methods. Based on the available data, the definition of NI was then classified depending on the historical evidence.

For the simulation study, only trials with sufficiently detailed reporting were considered. The revised Cochrane risk-of-bias tool for randomized trials (RoB2) was used to assess the risk of bias in randomized trials.^22^ As this search did not cover a specific disease area, publication bias was not assessed.

### Statistical analysis

Descriptive statistics were used to summarize the included trials, including the percentage and the number of studies reporting all identified items. If sufficient data were available from an included study, assumptions about how the NI margin was estimated or how the CI was calculated were changed to simulate the effect of alternative approaches. In addition, the analyzed population and handling of missing data or dropouts, respectively, were modified.

The historical effect size of the active comparator was reappraised based on the available literature. No attempt was made to systematically identify all the historical evidence for this part of the analysis due to limited resources to conduct further systematic reviews for every included RCT.

Alternative methods for defining the NI margin were carried out as described by Althunian and colleagues.^5^ The preserved fraction was set to 50% for the fixed-margin method and the following formula was used to calculate the NI margin (M2).

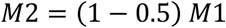

M1 is the estimate of the PE from the literature. The alternative methods used to calculate the CI for proportions were Wald, Wald continuity corrected, Wald interval with an adjustment according to Agresti and Caffo, Miettinen-Nurminen (MN), Newcombe or Wilson score.^23–26^ Sample size estimations were calculated based on the formula for an NI trial comparing two proportions.^27^ Only binary endpoints were included to further explore the effect of dropouts.

A post hoc comparison of the standardized mean difference of the NI margin by endpoint was conducted (eTable 2). In addition, trial conclusions were compared based on the number of dropouts for the PE and the number of crossovers.

All analyses were performed using R Statistical Software (v4.2.3; R Core Team 2023) and the packages “tidyverse” and “DescTools”.^28–30^

## RESULTS

After excluding 10’944 studies, 101 RCTs and 44’370 patients were included (Figure 1, eTable 3). RoB is shown in the eTable 4. A total of 81 (80.2%) trials stated a stated a rationale for the NI design, while 20 trials (19.8%) did not. The majority of studies defined the NI margin as an absolute effect (n=83, 82.2%). Six studies (5.9%) did not define the NI margin. Previous literature for defining the NI margin was cited in 44 (43.6%) trials and was mostly observational (Table 2). In six (5.9%) trials, the authors referred to regulatory guidance for drug trials by authorities.

**Figure 1.**
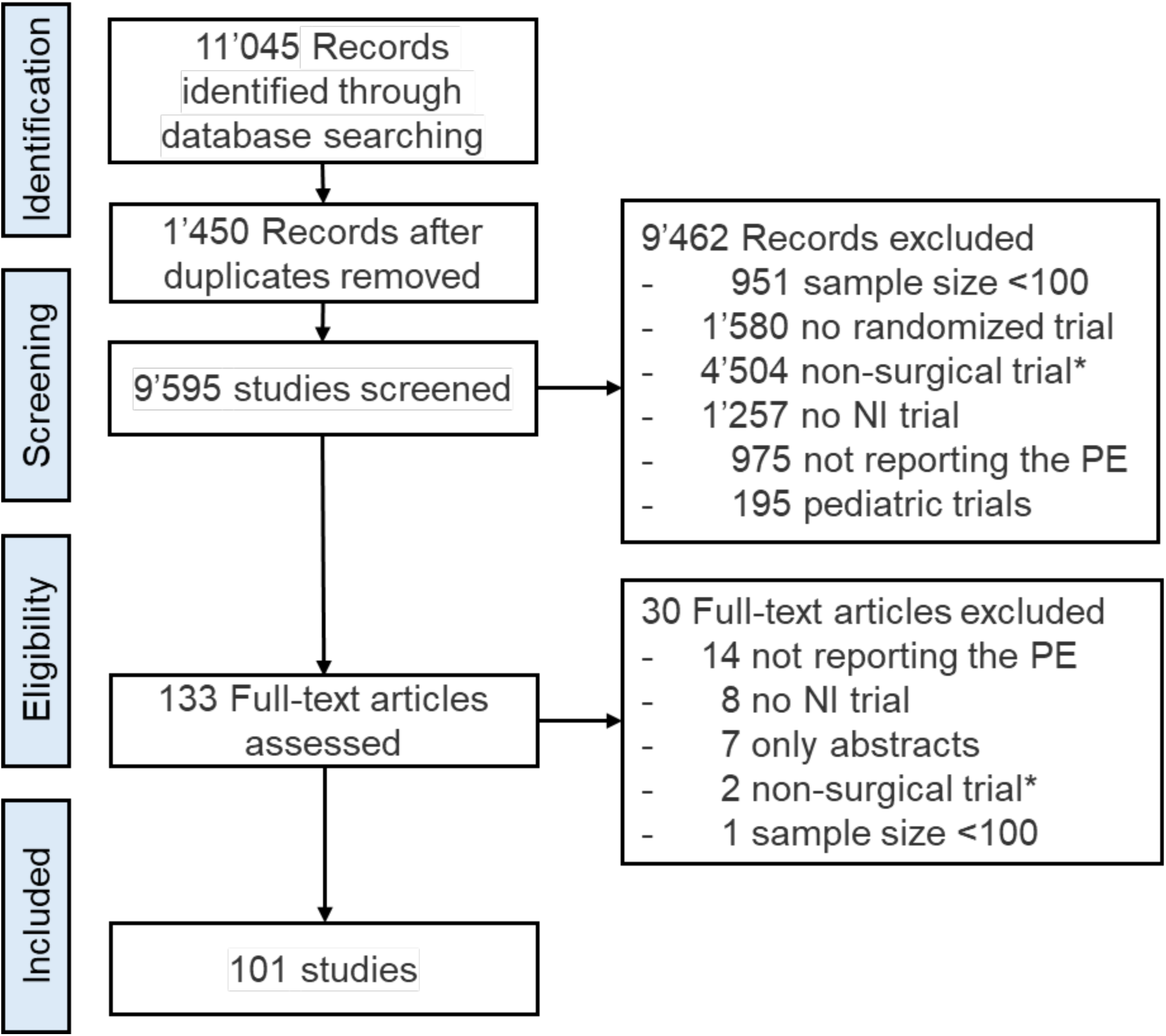
PRISMA Flow diagram. *includes non-abdominal surgical trials

**Table 1.** Reporting according to the CONSORT Extension for NI trials.

| Item | n (%) |
| --- | --- |
| Identification as NI RCT |  |
| Title | 39 (38.6) |
| Abstract | 59 (58.4) |
| Not specified | 3 (3.0) |
| Rational | 81 (80.2) |
| Superiority | 12 (11.9) |
| Interim analysis |  |
| 1 | 18 (17.8) |
| ≥2 | 9 (8.8) |
| Method for CI |  |
| Not specified | 87 (86.1) |
| Newcombe | 5 (5.0) |
| Wald | 2 (2.0) |
| Dunnett-Gent | 2 (2.0) |
| Miettinen–Nurminen | 2 (2.0) |
| Bootstrapping | 1 (1.0) |
| Wilson’s Score | 1 (1.0) |
| Resampling | 1 (1.0) |
| CI sided |  |
| 1-sided | 67 (66.3) |
| 2-sided | 17 (16.8) |
| Figure | 17 (17.2) |
| Conclusion |  |
| Non-inferior | 72 (71.3) |
| Not non-inferior | 21 (20.8) |
| Inconclusive | 2 (2.0) |
| Comparable | 1 (1.0) |
| No difference | 1 (1.0) |
| No conclusion (low power) | 1 (1.0) |
| Not non-inferior (ITT), non-inferior (PP) | 1 (1.0) |
| Not superior | 1 (1.0) |
| Safe and feasible | 1 (1.0) |
| Conclusion-based findings |  |
| Non-inferior | 64 (63.4) |
| Not non-inferior | 22 (21.8) |
| No conclusion on non-inferiority | 15 (14.9) |
| Conclusion |  |
| Appropriate | 87 (86.1) |
| Insufficient results reported<sup>a</sup> | 12 (11.9) |
| Margin not predefined<sup>a</sup> | 6 (5.9) |
| Not based on CI | 1 (1.0) |
| No population defined | 1 (1.0) |
| Clinical judgement | 99 (98.0) |
| CI, confidence interval; ITT, intention-to-treat; NI, non-inferiority; PP, per protocol; RCT randomized controlled trial <sup>a</sup>six patients had no margin defined and insufficiently reported results |  |

**Table 2.** Description of what information was used to define NI margins in abdominal surgical RCTs.

| Method | n (%) |
| --- | --- |
| Previous literature<sup>a</sup> | 44 (43.6) |
| Meta-analysis | 5 (5.0) |
| SR | 1 (1.0) |
| 3 RCTs | 1 (1.0) |
| 2 RCTs | 8 (7.9) |
| 1 RCT | 17 (16.8) |
| Observational study | 21 (20.8) |
| No previous literature | 57 (56.4) |
| Unpublished data | 17 (16.8) |
| Previously used margins in RCTs | 3 (3.0) |
| Consensus | 2 (2.0) |
| Relevance for oncology trials | 1 (1.0) |
| Simulation | 1 (1.0) |
| Defined by clinicians and PRs | 1 (1.0) |
| Discrete choice experiment | 1 (1.0) |
| Not specified | 31 (30.7) |
| Authority guidance |  |
| FDA | 3 (3.0) |
| EMA | 2 (2.0) |
| PMDA | 1 (1.0) |
| EMA, European Medicines Agency; FDA, Food and Drug Administration; PDMA, Pharmaceuticals and Medical Devices Agency (Japan); PR, patient representatives; SR, systematic review |  |
| <sup>a</sup>Multiple entries possible |  |

The reporting of the primary outcome in relation to the NI margin was judged to be of good quality in most RCTs (n=79, 78.2%). Quantitatively, the NI margin allowed for non-inferiority, ranging from 10.4% to five times worse than the estimated outcome of the active comparator (Figure 2). Most RCTs chose a margin of 5% or 10% (Figure 3A). When stratified by type of PE, trials with quantitative PEs chose larger standardized margins (median 0.42 (IQR 0.38-0.49)) compared to binary PEs 0.26 (IQR 0.14-0.38) or time-to-event PEs 0.17 (IQR 0.14-0.40) (Figure 3B).

**Figure 2.**
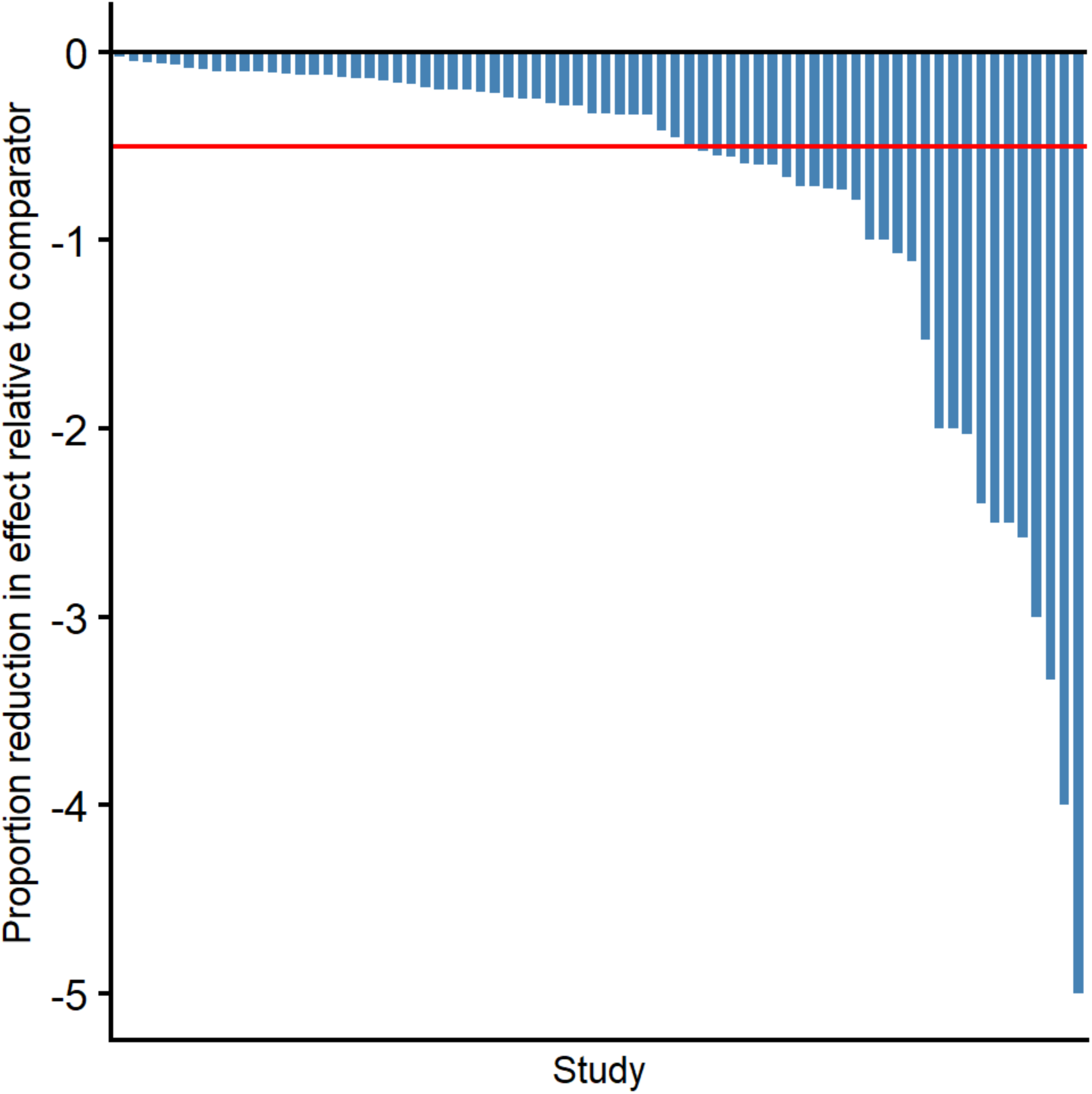
Accepted reduction in the proportion of the effect for the treatment based on the expected outcome for the primary endpoint. Included were trials specifying both an estimated outcome reference and an NI margin (n = 70). The red horizontal line indicates the 50% reduction as recommended by the FDA.

**Figure 3.**
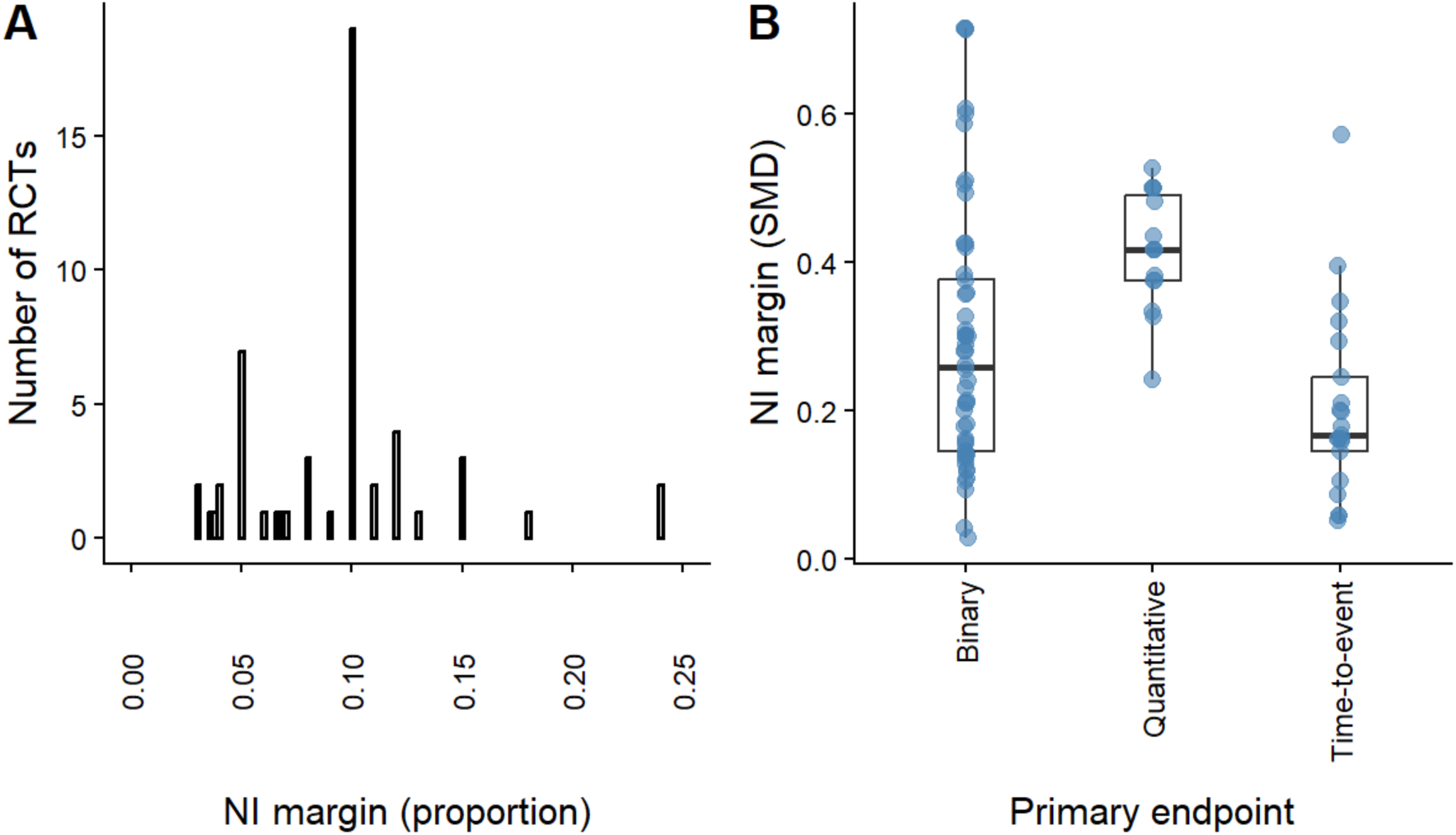
NI margin variation and impact of dropouts and crossover on trial conclusion. **A** Distribution of the NI margin (proportion). **B** Standardized NI margin stratified by type of endpoint.

**Figure 4.**
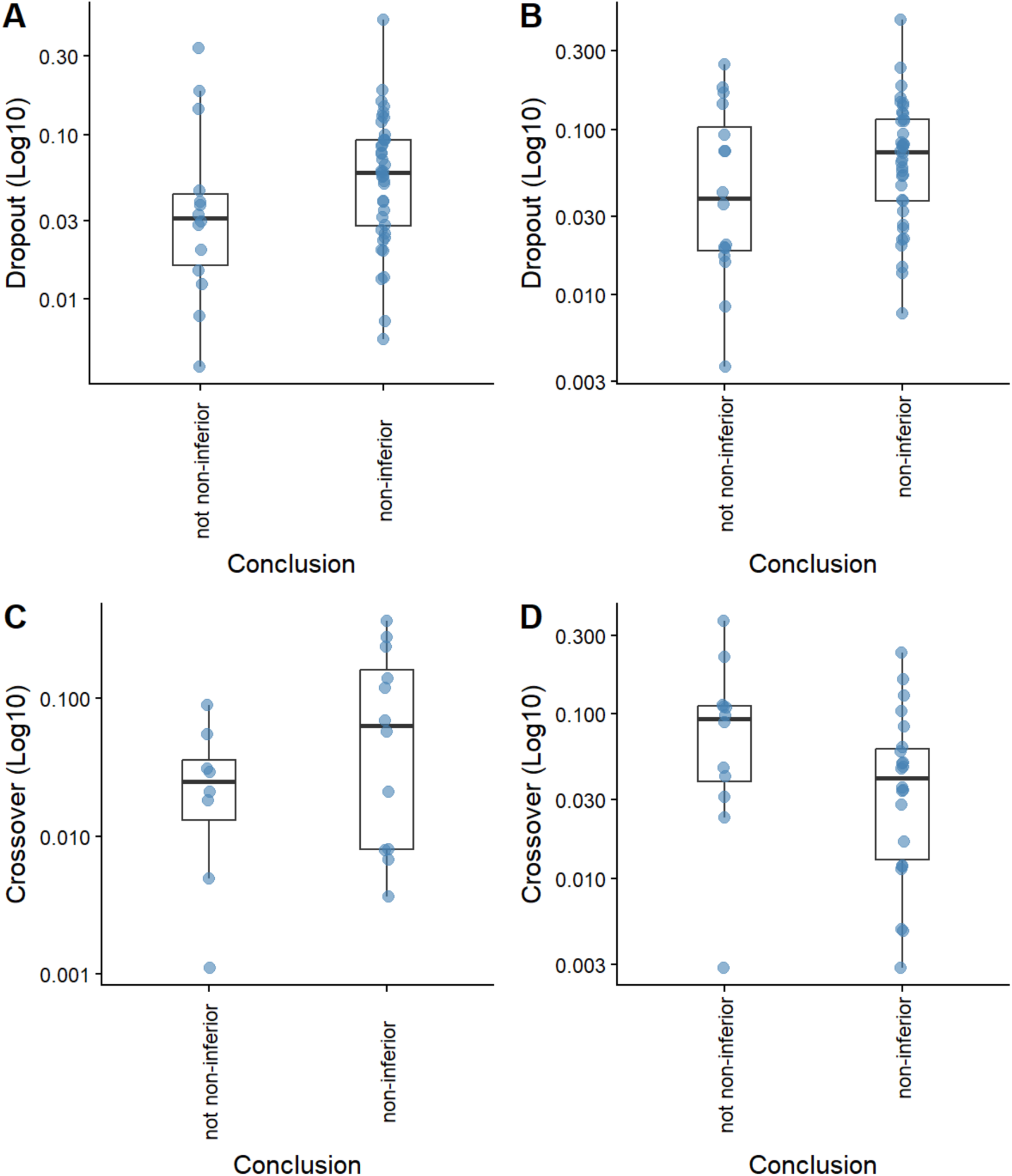
Dropouts and crossover in the analyzed population. Number of dropouts for the PE in the treatment (A) and the comparator arm (B). Number of crossovers from treatment to comparator (C) and from comparator to treatment (D)

### Definition of the NI margin

Among 44 studies that cited historical data, 35 (34.7%) adequately reported the PE. Thirty-four assessed an absolute risk difference and one assessed a hazard ratio as the PE. Thirty-three used a clinical fixed margin and two used the fixed-margin method. Twenty-four trials did not report a difference in expected outcomes based on the cited literature. Six trials based their difference on the treatment and the active comparator and one trial solely based its margin on the predicted PE event rate of the treatment arm.

The two RCTs using the fixed-effect method estimated the NI margin based on a historical comparison of the intervention and the active comparator.^31,32^ Although not entirely applicable, since the 50% preserved fraction guidance is originally based on placebo-controlled drug trials, one trial had a more conservative preserved fraction (>50%) from the active comparator, while the other had 0% preservation of the active comparator (eTable 5).^2,33^ Additionally, two RCTs used placebo-controlled historical RCTs.^34,35^ In those, the fixed-margin method is applicable and showed a lower NI margin than the clinical fixed margin actually used in both trials. Based on the published NI margin, 0% of the treatment effect was preserved, or it was even accepted to be worse than the placebo arm. Nevertheless, alternative NI margins did not alter the trial conclusion except in one trial where the NI margin was exceeded by 0.02%.

### Re-estimation of confidence intervals

Six trials reported all necessary variables for simulating alternative CI estimation methods (eTable 6). In none of the RCTs the conclusion was changed when the alternative CIs were compared with the predefined NI margin.

### Dropouts and crossover

The number of patients without data for the PE was not reported in 14 trials and 20 trials reported no missing data for the PE. Trials with dropouts more often concluded non-inferiority (Figure A/B). Similarly, trials with crossover to the comparator more often concluded non-inferiority, while crossover to the treatment more often led to the rejection of non-inferiority (Figure C/D).

Of the remaining 67 trials (66.3%), 23 (22.8%) had a time-to-event or quantitative PE. Sample sizes were estimated to be larger in trials with non-binary PEs with higher dropout and crossover rates (eTable 7). As a result, the per-protocol population treatment arm increased in size and more so in the trials with non-binary PEs. The number of patients in trials with a binary PE ranged from one to 387 (0.3% to 48.1% of the total sample size). Within-trial dropout rates ranged from 4.8% to 15% across trial arms. Zero to 214 (0% to 36.6%) patients were treated in the comparator arm rather than the treatment arm, and 0 to 331 (0% to 36.2%) crossed over to the treatment arm. Crossover was more frequent in the treatment arm (n = 25) than in the active comparator arm (n = 16).

Data for CI estimation and dropouts were complete for three trials. Li et al reported the MN method for CI calculation for the risk difference.^36^ The recalculated CI was identical when using the Wald method (-0.93 to 6.0). Since there was no crossover reported, the CI was calculated by assuming the same event rate if only 150 patients, as per the sample size calculation. The CI was -0.98% to 6.3% using the Wald method and −2.3% to 9.2% using the MN method. Both recalculated 95% CIs were below the prespecified NI margin of 10%, allowing up to 15% recurrence rate after 33 months in this trial. Assuming that the reference treatment had the estimated 5% (n ≈ 4) recurrence rate, the control had the same rate and number minus 2, the CI was -3.6% to 8.9% and -4.6% to 10.7% using the Wald and the MN method, respectively. The latter is above the NI margin and would therefore be inconclusive and non-inferiority therefore rejected. The sample size was recalculated based on the normal (Wald) approximation sample size formula for a two-arm non-inferiority trial on the absolute risk difference of two proportions that was provided by the authors. However, the event rate was set to 2.66%, the actual event rate in the control. The resulting sample size was 82 overall, smaller than the one based on the 5% event rate.

Maarten and colleagues reported the results of a trial underpowered by 2 patients for the intention-to-treat (ITT) population; the NI margin was -7%.^37^ Dropouts were balanced (18 and 19 patients in the treatment and the active comparator arm, respectively) and no crossover was reported. Using the corrected Wald method, the CI was -7.2% to 14.6%. Since the CI could not be reconstructed using the reported method (Wilson score), but only the Wald method, these findings were not further explored. Since this study used a 90% CI for the estimate of the difference between the primary endpoint, a 95% CI was alternatively chosen. All methods resulted in a lower CI bound below -7%. The authors estimated the treatment to be superior to the active comparator. Therefore, no further sample size recalculations based on the actual event rates were conducted.

Patel et al published results with an ITT including 142 patients and a sample size of 142 patients.^38^ No drop-out or crossover was reported. A margin of 15% was defined and the event rate was estimated to be 15% in both arms. Reestimation of the sample size based on the true active comparator rate, 9.9%, resulted in a much larger sample size of 161 per arm or 322 patients. The now underpowered study would result in a larger CI or a more conservative estimate. The authors did not mention the limitation of the lower event rate.

## DISCUSSION

In summary, surgical NI trials often used clinically fixed NI margins due to the lack of placebo-controlled trials. Historical evidence is cited in over half of trials but often disregarded when setting the NI margin. NI margins accept outcomes for the PE that are sometimes worse than historical effects. Poor reporting prevents reproducibility as well as meaningful comparisons across trials and thereby undermines the ability to conclude on NI.

Surgical trials are notoriously challenging to conduct due to the high expense, difficulties in recruitment, strong surgeon and patient preferences and little commercial incentive since RCTs are not required for regulatory approval.^39^ Frameworks to aid researchers in the conduct of RCTs are traditionally tailored to drug trials. For example, regulatory guidance may require a placebo-controlled superiority trial to define the preserved fraction of the active comparator in NI trials.^2,40^ Unlike drug trials, placebo-controlled historical evidence rarely exists in surgical NI trials unless the intervention involves a pharmacological treatment. In this review, we identified only two NI trials which based their margin on placebo-controlled historical evidence, and both assessed the effect of disinfectants. Both used the fixed-margin method to calculate the NI margin and allowed for a less preserved fraction than the recommended 50%.^2^

Thus, surgical investigators are forced to switch to alternative methods for defining the NI margin. These often involve historical non-randomized comparisons of the treatment with the active comparator. In our study, most trials only had data available from ≥1 historical comparison of the treatment and the active comparator. Understandably, no NI margin estimation similar to drug trials is feasible in this scenario. Moreover, estimating a preserved fraction is similarly limited, as no data on the course of the disease without surgery is available. Hence, NI margin definitions require thorough clinical judgement, but also a detailed and transparent reporting on how the NI margin was derived. However, for some types of interventions, e.g., minimally invasive versus conventional approaches, recommendations similar to the FDA or EMA guidance may be achievable and helpful. The focus should be avoiding overly tolerant margins since endpoints, e.g. complications, have generally serious consequences for patients.

While simulation studies were only feasible for six RCTs due to poor reporting, they illustrated the fragility of the assumptions and how this altered trial conclusions when the NI margin, the CI, or the sample size was changed. Moreover, dropouts for the PE and crossover often led to dilution of effects and frequently promoted conclusions of non-inferiority. Investigators should be aware of and acknowledge their assumptions and associated variations during trial conduct and ensure detailed reporting to allow interpretation of results. Considering the scarce prior information during the trial design phase, the clinically acceptable range of the PE in surgical trials and hence the clinicians’ expertise becomes even more important. Notably, our simulation studies were not intended to criticize the original methods used by the trialists, but rather demonstrate the impact of different assumption on statistical results and clinical conclusions. These alternative scenarios demonstrate the impact of these assumptions and how fragile the design and conclusion of NI trials can be. However, similar to a previous study which focused on CI reporting in antibiotic NI trials, our study indicates that while the reporting is generally poor, the impact of different methods on trial conclusions is limited.^41^

This research has several limitations which are important and need to be interpreted in the context of surgical research. First, we did not conduct a similarity check of the patient population with previous RCTs in which the efficacy of the reference treatment was established as only a few previous comparable trials were available. Similarly, intervention congruency was not evaluated for the same reason and additionally because standardization in many surgical interventions is often not achievable. Reasons are different stages of the surgeon in the learning curve, heterogeneity in surgeon and center expertise, or evolving or alternative perioperative management standards.^42,43^ Moreover, while all available sources were searched, including registry entries, protocols and appendices, some information may have been missed. Only after this effort were data points classified as missing. This approach may not be feasible when information is thought to offer the best possible treatment of patients and only trial reports are consulted. Hence, reporting is rather overestimated in this review when compared to a real-world setting. It can only be emphasized how important the information in original trial reports and the statistical as well as the peer-reviewing process are to improve reporting quality. A minimum sample size of ≥100 was chosen per treatment arm, which might have missed some relevant trials. However, sample sizes <100 are more frequently associated with wider CI when different CI calculation methods are applied and therefore limited due to model instability.^41^

Furthermore, the constancy assumption was only rarely acknowledged. However, compared to the unalterable drug application, many technical aspects vary from surgeon to surgeon. Hence, the same procedure performed by different surgeons may result in alternative outcomes even if carried out at the same time or at a different time. However, like in drug trials, there can still be an improvement in general patient management, which would result in a violation of the constancy assumption. This can be compared to a surgeon or center learning curve during which the surgeon tends to have better outcomes during later stages of the learning curve.^42,43^

Based on the findings of this study, guidance on surgical-specific NI margin definitions seems necessary. First, the NI design has to be clearly justified as the clinical aspect of the margin definition is disproportionally more relevant compared to drug trials and relies on clinicians’ expertise. Where available, meta-analytic historical effect estimates should be taken into account. Measures that can be taken to reduce the impact of vague a priori information, like adaptive trial elements, should be anticipated and clearly defined early during trial design. Better reporting of all design elements and analytic methods, including the CI estimation and the handling of missing data, is indispensable.

## Conclusion

Altogether, with little guidance and limited prior information during the design phase, the conduct of NI trials in surgical specialties is a formidable task. NI margins are often clinically fixed due to a lack of previous literature, particularly placebo-controlled trials or information on the natural history of the disease. Many NI margins permit worse outcomes in the treatment arm, limiting interpretability. Based on findings with the drawback of design limitations, the conclusions of NI trials are, however, generally reasonable and take into account the benefits and risks to patients, sustaining the non-maleficence principle. Strengthening methodological standards and reporting requirements is necessary to improve the credibility of non-inferiority trial findings in abdominal surgery.

## Supporting information

Online Only Supplement

## Data Availability

All data produced in the present study are available upon reasonable request to the authors

## Acknowledgement

We thank Dr Marshall Dozier, Lead for Library Academic Support, who helped develop the search strategy and identify the information sources.

## PRISMA Checklist 2020

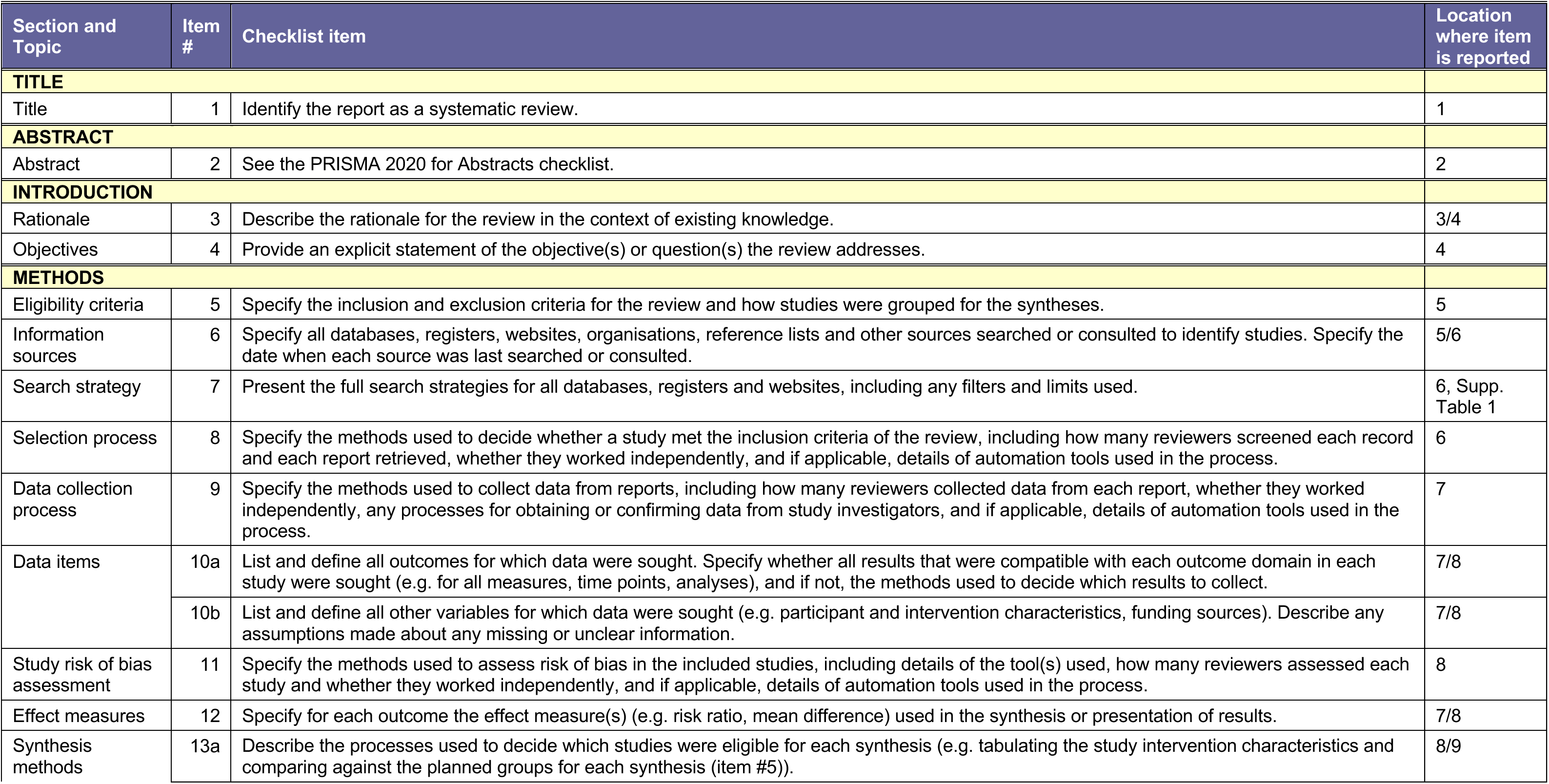
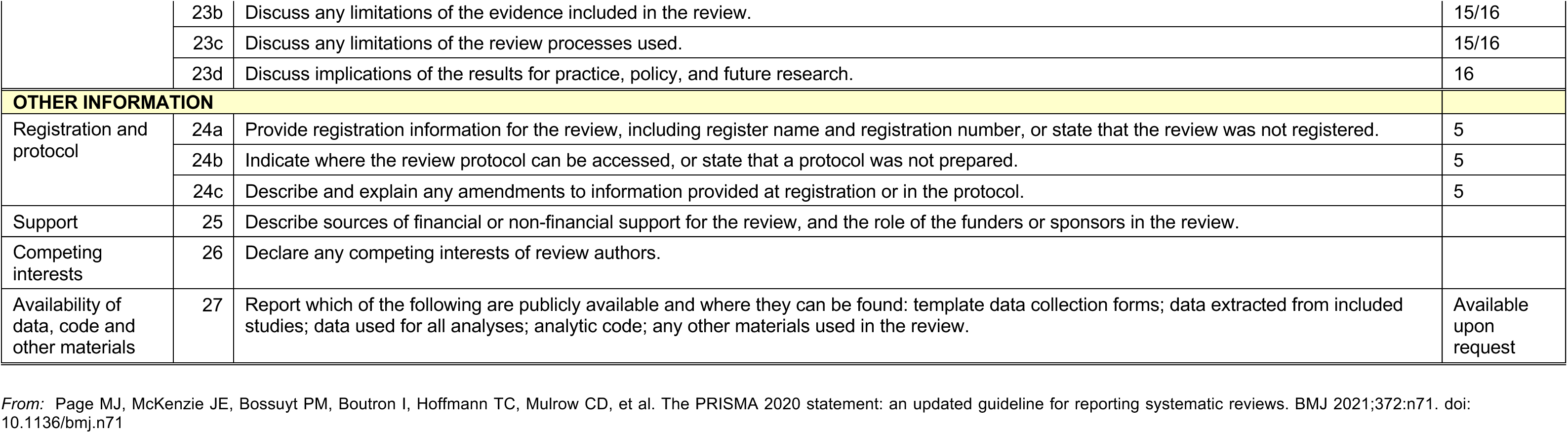
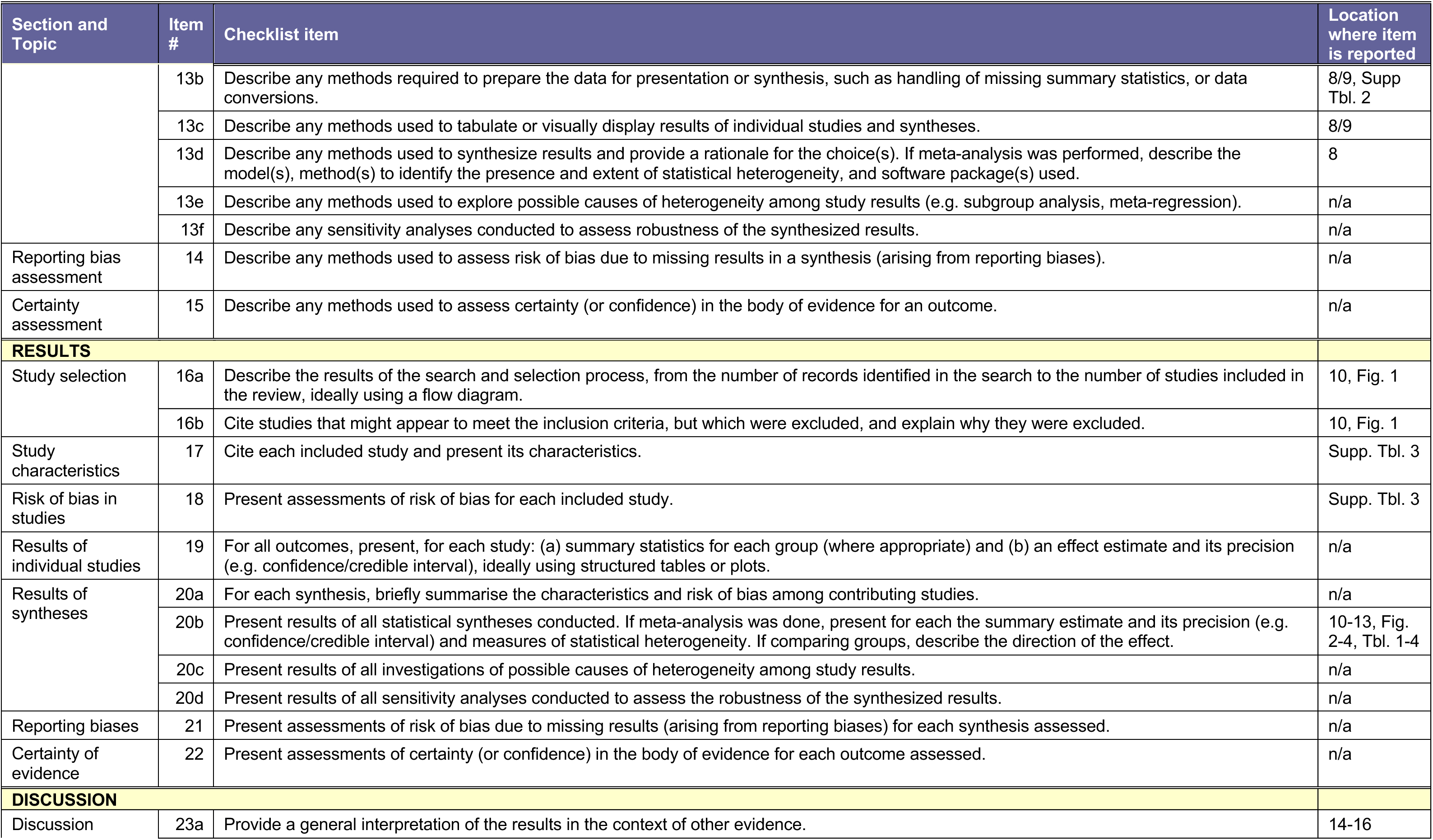

