## Supplementary material for "Non-Inferiority Margins in Randomized Controlled Trials in Abdominal Surgery – a Systematic Review": Online Only Supplement

**Online-Only Supplements**

**eTables**

eTable 1 Search strings for all database searches

| **Database** | **Search string** |
| --- | --- |
| Ovid Medline | 1  Randomized Controlled Trials as Topic/ or randomized controlled trial/ or Random Allocation/ or Double Blind Method/ or Single Blind Method/ or clinical trial/ or clinical trial, phase iii.pt. or clinical trial, phase iv.pt. or controlled clinical trial.pt. or randomized controlled trial.pt. or multicenter study.pt. or clinical trial.pt. or exp Clinical Trials as topic/ or ((clinical adj trial$) or ((singl$ or doubl$ or treb$ or tripl$) adj (blind$3 or mask$3))).tw. or PLACEBOS/ or placebo$.tw. or randomly allocated.tw. or (allocated adj2 random$).tw.  2  case report.tw. or letter/ or historical article/ or exp comment/ or exp editorial/ or exp published erratum/ or Randomized Controlled Trial, Veterinary/ or Clinical Trial, Veterinary/ or Surgery, Veterinary/ or Veterinary Medicine/ or protocol.ti.  3  (non-inferior* or noninferior* or equivalence or equivalent or "not inferior*").mp.  4  surgical procedures, operative/ or cautery/ or cryosurgery/ or endometrial ablation techniques/ or angioplasty, laser/ or corneal surgery, laser/ or maze procedure/ or radiofrequency ablation/ or ambulatory surgical procedures/ or anastomosis, surgical/ or anterior temporal lobectomy/ or bariatric surgery/ or conization/ or sentinel lymph node biopsy/ or "bloodless medical and surgical procedures"/ or circumcision, female/ or circumcision, male/ or cardiovascular surgical procedures/ or curettage/ or cytoreduction surgical procedures/ or debridement/ or decompression, surgical/ or device removal/ or digestive system surgical procedures/ or dissection/ or drainage/ or elective surgical procedures/ or electrosurgery/ or endocrine surgical procedures/ or chemotherapy, cancer, regional perfusion/ or heart bypass, left/ or fasciotomy/ or hemostasis, surgical/ or keratectomy/ or laparotomy/ or ligation/ or lymph node excision/ or mastectomy/ or metastasectomy/ or microsurgery/ or minimally invasive surgical procedures/ or minor surgical procedures/ or monitoring, intraoperative/ or myotomy/ or obstetric surgical procedures/ or neurosurgical procedures/ or ophthalmologic surgical procedures/ or oral surgical procedures/ or orthopedic procedures/ or ostomy/ or otorhinolaryngologic surgical procedures/ or pelvic exenteration/ or perioperative care/ or perioperative period/ or plastic surgery procedures/ or pneumonectomy/ or prophylactic surgical procedures/ or prosthesis implantation/ or reoperation/ or second-look surgery/ or splenectomy/ or surgery, computer-assisted/ or symphysiotomy/ or thoracic surgical procedures/ or transplantation/ or urogenital surgical procedures/ or wound closure techniques/ or cementation/ or immobilization/ or ischemic preconditioning/ or neoplasm transplantation/ or neuromuscular blockade/ or telemetry/ or trauma severity indices/ or "video-assisted techniques and procedures"/ or "equipment and supplies"/ or exp Orthopedic Procedures/ or exp Specialties, Surgical/  5  1 AND 3 AND 4  6  5 NOT 2  7  limit 6 to dt=20060401-20251219  8  limit 7 to "all adult (19 plus years)" |
| EMBASE | 1  Clinical Trial/ or Randomized Controlled Trial/ or controlled clinical trial/ or multicenter study/ or Phase 3 clinical trial/ or Phase 4 clinical trial/ or exp RANDOMIZATION/ or Single Blind Procedure/ or Double Blind Procedure/ or Crossover Procedure/ or PLACEBO/ or randomi?ed controlled trial$:ti,ab.mp. or rct:ti,ab.mp. or random$:ti,ab adj2:ti,ab allocat$:ti,ab.mp. or single blind$:ti,ab.mp. or double blind$:ti,ab.mp. or ((treble or triple) adj blind$:ti,ab).mp. or placebo$:ti,ab.mp. or Prospective Study/ [mp=title, book title, abstract, original title, name of substance word, subject heading word, floating sub-heading word, keyword heading word, organism supplementary concept word, protocol supplementary concept word, rare disease supplementary concept word, unique identifier, synonyms, population supplementary concept word, anatomy supplementary concept word]  2  Case Study/ or case report.tw. or abstract report/ or letter/ or Conference proceeding.pt. or Conference abstract.pt. or Editorial.pt. or Letter.pt. or Note.pt. or historical article/ or exp comment/ or exp editorial/ or exp published erratum/ or Randomized Controlled Trial, Veterinary/ or Clinical Trial, Veterinary/ or Surgery, Veterinary/ or Veterinary Medicine/ or protocol.ti.  3  (non-inferior* or noninferior* or equivalence or equivalent or "not inferior*").mp.  4  exp coronary artery bypass surgery/ or exp retina surgery/ or exp microvascular surgery/ or exp heart valve surgery/ or exp pituitary surgery/ or exp off pump coronary surgery/ or exp breast surgery/ or exp refractive surgery/ or exp brain surgery/ or exp face surgery/ or exp liver surgery/ or exp colorectal surgery/ or exp urethra surgery/ or exp mitral valve surgery/ or exp cancer surgery/ or exp urologic surgery/ or exp meniscal surgery/ or exp thymus surgery/ or exp thoracic aortic surgery/ or exp Obesity Surgery Mortality Risk Score/ or exp corneal surgery/ or exp cytoreductive surgery/ or exp ascending aorta surgery/ or exp laparoendoscopic single site surgery/ or exp minimally invasive surgery/ or exp open surgery monopolar electrosurgical electrode/ or exp transvaginal natural orifice transluminal endoscopic surgery/ or exp geriatric surgery/ or exp computer assisted surgery system/ or exp spinal surgery equipment/ or exp "patient history of orthopedic surgery"/ or exp femtosecond laser-assisted cataract surgery/ or exp bladder surgery/ or exp trachea surgery/ or exp laser refractive surgery/ or exp spinal cord surgery/ or exp focused ultrasound surgery/ or exp intestine surgery/ or exp nose surgery/ or exp knee ligament surgery/ or exp major surgery/ or exp gastric bypass surgery/ or exp gynecologic surgery/ or exp uterus surgery/ or exp spine surgery/ or exp robotic hip surgery system/ or exp breast-conserving surgery/ or exp ambulatory surgery/ or exp orthopedic surgery/ or exp tendon surgery/ or exp vein surgery/ or exp skin surgery/ or exp "bipolar electrosurgical electrode (open surgery)"/ or exp "patient history of cataract surgery"/ or exp "head and neck surgery"/ or exp vitreoretinal surgery/ or surgery/ or exp endovascular surgery/ or exp cerebrovascular surgery/ or exp spleen surgery/ or exp open heart surgery/ or exp rectum surgery/ or exp robotic surgery simulator/ or exp laparoscopic surgery/ or exp preprosthetic surgery/ or exp transanal endoscopic surgery/ or exp abdominal surgery/ or exp carotid artery surgery/ or exp Mohs micrographic surgery/ or exp retina detachment surgery/ or exp ureter surgery/ or exp endoscopic sinus surgery/ or exp aortic surgery/ or exp nephron sparing surgery/ or exp endocrine surgery/ or exp pelvis surgery/ or exp anus surgery/ or exp video assisted surgery/ or exp coronary artery surgery/ or exp gastrointestinal surgery/ or exp cardiac surgery intensive care unit/ or exp "aortic root surgery"/ or exp larynx surgery/ or exp colon surgery/ or exp plastic surgery/ or exp nerve surgery/ or exp urinary tract surgery/ or exp hip surgery/ or exp joint surgery/ or exp foot surgery/ or exp minimally invasive cardiac surgery/ or exp off pump surgery/ or exp arthroscopic surgery/ or exp "patient history of dental surgery"/ or exp thyroid surgery/ or exp stereotaxic surgery/ or exp middle ear surgery/ or exp vascular surgery/ or exp oral surgery/ or exp endoscopic surgery/ or exp male genital system surgery/ or exp knee surgery/ or exp shoulder stabilization surgery/ or exp thorax surgery/ or exp reconstructive surgery/ or exp enhanced recovery after surgery/ or exp ear nose throat surgery/ or exp general surgery/ or exp natural orifice transluminal endoscopic surgery/ or exp elective surgery/ or exp bariatric surgery/ or exp esthetic surgery/ or exp wrist surgery/ or exp facial nerve surgery/ or exp glaucoma surgery/ or exp endoscopic endonasal surgery/ or exp bypass surgery/ or exp eye surgery/ or exp heart surgery/ or exp hand surgery/ or exp adrenal surgery/ or exp vagina surgery/ or exp maxillofacial surgery/ or exp pediatric surgery/ or exp skull surgery/ or exp conversion to open surgery/ or exp stapes surgery/ or exp strabismus surgery/ or exp newborn surgery/ or exp coagulation surgery/ or exp robot assisted surgery/ or exp computer assisted surgery/ or exp plastic surgery implant/ or exp ear surgery/ or exp kidney surgery/ or exp ligament surgery/ or exp shoulder surgery/ or exp transsphenoidal surgery/ or exp fetus surgery/ or exp throat surgery/ or exp robotic knee surgery system/ or exp transoral robotic surgery/ or exp experimental surgery/ or exp prostate surgery/ or exp stomach surgery/ or exp cardiovascular surgery/ or exp emergency surgery/ or exp ultrasound surgery/ or exp endoscopic pituitary surgery/ or exp biliary tract surgery/ or exp open surgery/ or exp aortic arch surgery/ or exp lung surgery/ or exp aneurysm surgery/ or exp decompression surgery/ or exp minor surgery/ or exp artery surgery/ or exp esophagus surgery/ or exp pancreas surgery/ or exp descending aortic surgery/ or exp ankle surgery/ or exp craniofacial surgery/ or exp second look surgery/ or exp uterine tube surgery/ or exp veterinary surgery/ or exp video assisted thoracoscopic surgery/  5  surgical technique/ or exp adhesiolysis/ or exp anastomosis/ or exp bypass surgery/ or exp cauterization/ or exp cerclage/ or exp chemosurgery/ or exp coagulation surgery/ or exp commissurotomy/ or exp curettage/ or exp device removal/ or exp dissection/ or exp electrosurgery/ or exp endoscopic surgery/ or exp excision/ or exp fenestration/ or exp implantation/ or exp incision/ or exp laser surgery/ or exp ligation/ or exp lithotomy/ or exp lobectomy/ or exp metastasis resection/ or exp morcellation/ or exp myotomy/ or exp ostomy/ or exp paracentesis/ or exp phlebotomy/ or exp radical resection/ or exp radiosurgery/ or exp recanalization/ or exp "skeletonization (surgical)"/ or exp sphincteroplasty/ or exp sphincterotomy/ or exp surgical drainage/ or exp surgical hemostasis/ or exp tractotomy/ or exp transoral robotic surgery/ or exp trephination/ or exp ultrasound surgery/ or exp vascular access/ or exp wedge resection/ or exp wound care/  6  1 AND 3  7  4 OR 5  8  6 AND 7  9  8 NOT 2  10  limit 9 to (human and yr="2006 -Current" and (article or article in press) and (adult <18 to 64 years> or aged <65+ years>)) |
| CENTRAL | 1  ((non-inferior* or noninferior* or equivalence or equivalent or "not inferior*").mp.):ti,ab,kw AND (surgical procedures):ti,ab,kw (Word variations have been searched)  2  Specialty, surgical OR Surgical Procedures, Operative  3  1 OR 2 |

eTable 2 Method of standardization of NI margins across all outcomes using the standardized effect size Cohen’s d.

| **NI margin** | **Transformation** |
| --- | --- |
| Hazard ratio (HR) | $d=\frac{\ln(\text{HR})}{1.81}$ |
| Risk difference (RD) | $d=\frac{RD}{\sqrt{p(1-p)}}$ where$p=\frac{p_{1}+p_{0}}{2}$  $p$ = pooled risk |
| Continuous scale | $d=\frac{\text{NI margin}}{\text{SD of the scale}}$  SD = standard deviation |

eTable 3 Risk of bias assessment

| First author | **Randomization process** | **Deviations from intended interventions** | **Missing outcome data** | **Measurement of the outcome** | **Selection of the reported result** | **Overall risk of bias** |
| --- | --- | --- | --- | --- | --- | --- |
| Akagi T^1^ | low risk | low risk | high risk | some concerns | high risk | high risk |
| Aniruthan D^2^ | some concerns | high risk | low risk | some concerns | some concerns | high risk |
| Arai S^3^ | low risk | low risk | low risk | low risk | low risk | low risk |
| Arezzo A^4^ | low risk | low risk | high risk | some concerns | some concerns | high risk |
| Arita J^5^ | low risk | low risk | low risk | some concerns | low risk | some concerns |
| Arumugaswamy PR^6^ | low risk | low risk | low risk | low risk | low risk | low risk |
| Ashley T^7^ | low risk | low risk | some concerns | some concerns | low risk | some concerns |
| Banos, PAP^8^ | low risk | low risk | high risk | high risk | high risk | high risk |
| Barendse RM^9^ | some concerns | low risk | low risk | low risk | low risk | some concerns |
| Broach RB^10^ | low risk | low risk | low risk | low risk | low risk | low risk |
| By-Band-Sleeve^11^ | low risk | low risk | some concerns | low risk | low risk | some concerns |
| Chang SKY^12^ | some concerns | low risk | low risk | low risk | some concerns | some concerns |
| COLOR Study Group^13^ | some concerns | low risk | low risk | low risk | low risk | some concerns |
| Contant CME^14^ | low risk | low risk | low risk | low risk | low risk | low risk |
| De Goede B^15^ | some concerns | low risk | high risk | low risk | some concerns | high risk |
| de Graaf N^16^ | low risk | low risk | some concerns | some concerns | low risk | some concerns |
| Degiuli M^17^ | some concerns | low risk | low risk | low risk | low risk | some concerns |
| Ellis RC^18^ | low risk | low risk | low risk | some concerns | some concerns | some concerns |
| Fleshman J^19^ | low risk | low risk | low risk | low risk | low risk | low risk |
| Fujita S^20^ | some concerns | low risk | low risk | low risk | low risk | some concerns |
| Goyal A^21^ | low risk | low risk | some concerns | high risk | high risk | high risk |
| Hedberg J^22^ | low risk | low risk | low risk | low risk | low risk | low risk |
| Hirao M^23^ | low risk | low risk | low risk | low risk | low risk | low risk |
| Horbach T^24^ | some concerns | high risk | high risk | high risk | low risk | high risk |
| Huyang WJ^25^ | some concerns | low risk | low risk | low risk | low risk | some concerns |
| Jalava K 2023^26^ | low risk | some concerns | low risk | low risk | low risk | some concerns |
| Jalava K 2025^27^ | low risk | low risk | low risk | low risk | low risk | low risk |
| Jeong SY^28^ | low risk | low risk | low risk | low risk | low risk | low risk |
| Jia XQ^29^ | some concerns | some concerns | low risk | high risk | some concerns | high risk |
| Jiang W^30^ | low risk | low risk | low risk | low risk | low risk | low risk |
| Kaiser J^31^ | low risk | low risk | low risk | low risk | low risk | low risk |
| Kambara Y^32^ | some concerns | low risk | low risk | high risk | low risk | high risk |
| Katai H^33^ | some concerns | low risk | low risk | low risk | some concerns | some concerns |
| Khaleel MI^34^ | high risk | some concerns | high risk | high risk | high risk | high risk |
| Kim YW^35^ | some concerns | low risk | low risk | low risk | low risk | some concerns |
| Kim HI^36^ | low risk | some concerns | low risk | low risk | some concerns | some concerns |
| Korrel M^37^ | low risk | low risk | low risk | low risk | low risk | low risk |
| Kwon YH^38^ | low risk | some concerns | some concerns | low risk | some concerns | some concerns |
| Langenbach MR^39^ | low risk | some concerns | low risk | low risk | low risk | some concerns |
| Lauscher JC^40^ | some concerns | some concerns | low risk | some concerns | some concerns | some concerns |
| Lee YS^41^ | low risk | low risk | low risk | some concerns | low risk | some concerns |
| Lehrskov LL^42^ | low risk | some concerns | high risk | high risk | low risk | high risk |
| Li G^43^ | low risk | some concerns | low risk | some concerns | low risk | some concerns |
| Li J^44^ | some concerns | some concerns | low risk | low risk | low risk | some concerns |
| Li S^45^ | some concerns | low risk | some concerns | low risk | low risk | some concerns |
| Lin HC^46^ | low risk | some concerns | some concerns | high risk | low risk | high risk |
| Lin W^47^ | low risk | high risk | some concerns | high risk | low risk | high risk |
| Lozada Hernandez EE^48^ | low risk | some concerns | some concerns | low risk | low risk | some concerns |
| Lu J^49^ | low risk | low risk | low risk | low risk | low risk | low risk |
| Mällinen J^50^ | some concerns | low risk | some concerns | low risk | low risk | some concerns |
| Mao Y^51^ | low risk | low risk | low risk | low risk | low risk | low risk |
| Meuzelaar RR^52^ | low risk | some concerns | low risk | some concerns | some concerns | some concerns |
| Mishra A^53^ | some concerns | low risk | low risk | low risk | some concerns | some concerns |
| Muduly DK^54^ | some concerns | low risk | low risk | low risk | low risk | some concerns |
| Okinaga K^55^ | some concerns | low risk | low risk | some concerns | low risk | some concerns |
| Oshikiri T^56^ | low risk | low risk | low risk | some concerns | low risk | some concerns |
| Park SE^57^ | low risk | low risk | low risk | low risk | low risk | low risk |
| Patel DN^58^ | high risk | low risk | low risk | low risk | low risk | high risk |
| Patel SV^59^ | low risk | some concerns | low risk | low risk | low risk | some concerns |
| Pedrazzani C^60^ | some concerns | low risk | low risk | high risk | low risk | high risk |
| Regimbeau JM^61^ | low risk | some concerns | some concerns | low risk | some concerns | some concerns |
| Robert M^62^ | low risk | some concerns | low risk | low risk | low risk | some concerns |
| Salminen P^63^ | low risk | low risk | low risk | some concerns | low risk | some concerns |
| Salmonsen Cb^64^ | low risk | low risk | low risk | some concerns | some concerns | some concerns |
| Sano T^65^ | some concerns | low risk | low risk | low risk | low risk | some concerns |
| Sasmal PK^66^ | some concerns | some concerns | low risk | low risk | some concerns | some concerns |
| Serra-Aracil X^67^ | low risk | low risk | high risk | low risk | high risk | high risk |
| Smith SR^68^ | low risk | low risk | low risk | low risk | low risk | low risk |
| Song Z^69^ | some concerns | low risk | low risk | some concerns | low risk | some concerns |
| Stevenson ARL^70^ | some concerns | low risk | low risk | some concerns | low risk | some concerns |
| Sun HB^71^ | some concerns | low risk | low risk | some concerns | low risk | some concerns |
| Sun Z^72^ | low risk | low risk | low risk | low risk | high risk | high risk |
| Suzuki T^73^ | some concerns | some concerns | low risk | some concerns | low risk | some concerns |
| Symeonidis S^74^ | some concerns | low risk | low risk | low risk | high risk | high risk |
| Tajima Y^75^ | some concerns | some concerns | low risk | some concerns | low risk | some concerns |
| Takeuchi H^76^ | low risk | low risk | low risk | some concerns | low risk | some concerns |
| Tanaka A^77^ | some concerns | low risk | low risk | low risk | low risk | some concerns |
| Ten Haaft BHEA^78^ | low risk | low risk | low risk | some concerns | low risk | some concerns |
| The CODA Collaborative^79^ | some concerns | some concerns | low risk | low risk | low risk | some concerns |
| Toyama H^80^ | some concerns | low risk | some concerns | low risk | low risk | some concerns |
| Tsukamoto S^81^ | some concerns | low risk | low risk | low risk | low risk | some concerns |
| Uchino M^82^ | some concerns | low risk | some concerns | low risk | low risk | some concerns |
| van Bodegraven EA^83^ | low risk | low risk | low risk | some concerns | low risk | some concerns |
| van der Lei S^84^ | low risk | low risk | low risk | some concerns | low risk | some concerns |
| van der Wilk BJ^85^ | some concerns | some concerns | low risk | low risk | low risk | some concerns |
| van Dijk AH^86^ | some concerns | some concerns | some concerns | low risk | some concerns | some concerns |
| Vons C^87^ | some concerns | low risk | some concerns | low risk | low risk | some concerns |
| Wang Q^88^ | some concerns | high risk | low risk | low risk | low risk | high risk |
| Wang Y^89^ | some concerns | high risk | low risk | some concerns | some concerns | some concerns |
| Watanabe J^90^ | low risk | low risk | low risk | low risk | some concerns | some concerns |
| Wei C^91^ | low risk | some concerns | low risk | some concerns | low risk | some concerns |
| Weindelmayer J^92^ | low risk | some concerns | low risk | some concerns | low risk | some concerns |
| Werner YB^93^ | low risk | low risk | low risk | low risk | low risk | low risk |
| Wu G^94^ | low risk | some concerns | low risk | low risk | some concerns | some concerns |
| Yang Y^95^ | low risk | some concerns | low risk | low risk | low risk | some concerns |
| Yoshida T^96^ | low risk | some concerns | low risk | low risk | some concerns | some concerns |
| Yu J^97^ | some concerns | low risk | low risk | low risk | low risk | some concerns |
| Yu Y^98^ | low risk | some concerns | some concerns | some concerns | low risk | some concerns |
| Zamkowski M^99^ | low risk | low risk | some concerns | some concerns | low risk | some concerns |
| Zeng Z^100^ | low risk | some concerns | low risk | low risk | low risk | low risk |
| Zhang K-H^101^ | low risk | some concerns | low risk | some concerns | some concerns | some concerns |

eTable 4 Aggregated study characteristics outside the CONSORT Extension for NI trials.

| **Topic** | **n (%)** |
| --- | --- |
| Journal |  |
| Surgical | 54 (54.5) |
| Other | 47 (45.5) |
| Funding |  |
| Governmental | 52 (51.5) |
| Industry | 14 (13.9) |
| Professional society | 8 (7.9) |
| Private foundation | 8 (7.9) |
| Hospital/university | 5 (5.0) |
| No external funding | 10 (9.9) |
| Not specified | 14 (13.9) |
| Sponsor |  |
| University (hospital) | 70 (69.3) |
| Non-academic hospital | 3 (3.0) |
| Collaborative research network | 8 (7.9) |
| Private hospital | 2 (2.0) |
| Industry | 4 (4.0) |
| Other | 2 (2.0) |
| Not specified | 12 (11.9) |
| Intervention |  |
| Different operation/stage | 16 (15.8) |
| Less invasive access | 16 (15.8) |
| Perioperative management | 12 (11.9) |
| Non-surgical treatment | 10 (9.9) |
| Less radical resection | 10 (9.9) |
| Pre or intraoperative drug application | 9 (8.9) |
| Restrictive drain policy | 7 (6.9) |
| Timing of intervention | 7 (6.9) |
| Less invasive | 7 (6.9) |
| Other | 3 (3.0) |
| Phase |  |
| 2 | 3 (3.0) |
| 3 | 25 (24.8) |
| 4 | 1 (1.0) |
| not specified | 72 (71.3) |
| Randomisation |  |
| Simple | 24 (23.8) |
| Stratified block randomisation | 22 (21.8) |
| Block randomisation | 21 (20.8) |
| Minimisation | 17 (16.8) |
| Stratified | 14 (13.9) |
| Minimisation, centre-stratified | 1 (1.0) |
| Stepped wedge cluster | 1 (1.0) |
| Not specified | 1 (1.0) |
| Blinding |  |
| Open-label | 52 (51.5) |
| Outcome assessor | 18 (17.8) |
| Patient | 7 (6.9) |
| Patient, outcome assessor | 6 (5.9) |
| Patient, analyst | 2 (2.0) |
| Patient, clinician | 1 (1.0) |
| Patient, investigator, analyst | 1 (1.0) |
| Single-blind, not further specified | 2 (2.0) |
| Not specified | 8 (7.9) |
| Purpose |  |
| Fewer adverse events | 20 (19.8) |
| Simpler procedure | 17 (16.8) |
| Faster recovery | 15 (14.9) |
| Avoid operation | 12 (11.9) |
| Easier application | 9 (8.9) |
| Avoid unnecessary treatment | 6 (5.9) |
| Less costly | 3 (3.0) |
| Availability | 2 (2.0) |
| Different procedure/operation | 2 (2.0) |
| Less trauma | 2 (2.0) |
| Other | 5 (5.0) |
| Not applicable | 2 (2.0) |
| Not specified | 6 (5.9) |
| Analysis population |  |
| ITT | 33 (32.7) |
| ITT, PP | 24 (23.8) |
| mITT | 7 ( 6.9) |
| mITT, PP | 10 ( 9.9) |
| mITT, PP, as-treated | 2 (2.0) |
| PP | 12 (11.9) |
| not specified | 13 (12.9) |

eTable 5 NI margins and preserved fractions of effect in surgical abdominal NI RCTs using originally reported parameters and 50% preserved fraction.

|  | Broach | Salminen | Park | Salmonsen |
| --- | --- | --- | --- | --- |
| PE reference, % | 20 | 99 | 15 | 20mg |
| PE treatment, % | - | 75 | 17 | 30mg |
| NI margin, % | -6.6 | 24 | 11 | 10mg |
| Preserved fraction, % (active comparator) | 52.5 | 0 | - | - |
| Preserved fraction, % (placebo) | - | - | -450 | 0 |
| Benefit, % | 33.9 * | 24 | 2 | 10mg |
| Fixed-margin, % (50% preserved ^102^) | -6.9 | 12 | 1 | 5mg |
| *based on the 20% event rate and a relative risk of 0.59: 0.2/0.59-0.2 | | | | |

eTable 6 Recalculations of the confidence interval (CI) for trials reporting the method for the CI calculation and for which the original CI could be calculated.

|  | **NI margin** | **CI** | **Reported CI** | **Method** | | | | |
| --- | --- | --- | --- | --- | --- | --- | --- | --- |
|  |  |  |  | Wald | Wald corrected | Agresti-Caffo | Miettinen-Nurminen | Newcombe |
| Patel | 15 | 95% | -8.3, 11.1 | -8.0, 10.8 | -9.4, 12.3 | -8.4, 11.2 | -8.8, 11.7 | -12.5, 9.6 |
| Li | 10 | 95% | -0.93, 6 | -0.93, 6.0 | -2.2, 7.2 | -2.3, 7.2 | -2.2, 8.7 | -3.6, 9.6 |
| Song | 10 | 95% | -13.9, 5.3 | -13.5, 5.0 | -14.6, 6.0 | -13.6, 5.2 | -14.0, 5.2 | -14.5, 5.9 |
| Maarten mITT | -7 | 90% | -6.2, 13.6 | -6.3, 13.7 | -7.2, 14.6 | -6.3, 13.6 | -6.3, 13.7 | -6.8, 14.2 |
| Maarten PP | -7 | 95% | -7.6, 13.9 | -7.6, 14.1 | -8.6, 15.2 | -7.6, 14.0 | -7.6, 14.1 | -8.3, 14.7 |
| Hedberg ITT | -9 | 95% | −14.4, 0.0 | -14.1, 0.2 | -14.6, 0.7 | -14.1, 0.3 | -14.3, 0.2 | -14.5, 0.6 |
| Hedberg PP | -9 | 95% | -19.1, 0.3 | -19.1, 3.4 | -19.8, 2.8 | -18.9, 3.1 | -19.1, 3.1 | -19.3, 2.5 |
| Van Bodegraven ITT | 8 | 95% | -13.8, 4.0 | -13.8, 4.0 | -14.5, 4.7 | -13.7, 4.1 | -13.9, 4.1 | -14.3, 4.6 |
| Van Bodegraven PP | 8 | 95% | -13.2, 5.0 | -13.2, 5.0 | -13.9, 5.7 | -13.1, 5.1 | -13.2, 5.2 | -13.6, 5.6 |
| *Values are not rounded, confidence intervals are two-sided. CI, confidence interval; NI, non-inferiority; PP, per protocol | | | | | | | | |

eTable 7 Comparison of trial sample size, populations, dropouts and events between trials that were further assessed because they chose a binary endpoint and trials with other endpoints that were not further explored. Analyzed population varied but only the largest was reported, e.g. ITT if both ITT and PP were reported. Since not all reported ITT and PP analyses, PP may have a larger number of patients.

|  | **Overall**  **median (IQR)** | **Non-binary PE**  **median (IQR)** | **Binary PE**  **median (IQR)** | **p** |
| --- | --- | --- | --- | --- |
| Sample size |  |  |  |  |
| W/o dropouts | 226 (132, 506) | 340 (11, 808) | 206 (134, 470) | 0.601 |
| Randomized |  |  |  |  |
| Treatment | 150 (89, 312) | 150 (81, 472) | 149 (91, 244) | 0.561 |
| Active comparator | 148 (83, 315) | 148 (79, 494) | 148 (85, 245) | 0.716 |
| ITT |  |  |  |  |
| Treatment | 149 (86, 262) | 148 (78, 469) | 150 (86, 240) | 0.665 |
| Active comparator | 147 (87, 274) | 142 (75, 491) | 150 (87, 235) | 0.917 |
| PP |  |  |  |  |
| Treatment | 136 (88, 215) | 170 (84, 424) | 135 (90, 198) | 0.672 |
| Active comparator | 112 (81, 221) | 117 (76, 418) | 122 (83, 202) | 0.897 |
| Dropouts for PE |  |  |  |  |
| Total | 4 (2, 8) | 6 (2, 10) | 4 (1, 8) | 0.300 |
| Treatment | 7 (2, 18) | 8 (3, 28) | 6 (1, 15) | 0.272 |
| Active comparator | 8 (3, 19) | 8 (4, 25) | 8 (2, 18) | 0.363 |
| Treatment, % | 3.9 (1.7, 9.0) | 5.4 (3.2, 8.7) | 3.3 (1.2, 8.7) | 0.147 |
| Active comparator, % | 5.9 (2.0, 11.3) | 7.4 (3.9, 11.7) | 4.6 (1.6, 11.3) | 0.306 |
| Crossover |  |  |  |  |
| To active comparator | 2 (0, 16) | 4 (1, 18) | 1 (0, 14) | 0.316 |
| To Treatment | 0 (0, 8) | 1 (0, 9) | 0 (0, 5) | 0.184 |
| IQR, interquartile range; ITT, intention-to-treat; PE, primary endpoint; PP, per protocol | | | | |
